# Step-by-Step Approach to Design Image Classifiers in AI: An Exemplary Application of the CNN Architecture for Breast Cancer Diagnosis

**DOI:** 10.1101/2025.06.17.25329727

**Authors:** Ahamadullah Lohani, Bhupesh Kumar Mishra, Kenneth Y. Wertheim, Temitayo Matthew Fagbola

## Abstract

In recent years, different Convolutional Neural Networks (CNNs) approaches have been applied for image classification in general and specific problems such as breast cancer diagnosis, but there is no standardising approach to facilitate comparison and synergy. This paper attempts a step-by-step approach to standardise a common application of image classification with the specific problem of classifying breast ultrasound images for breast cancer diagnosis as an illustrative example. In this study, three distinct datasets: Breast Ultrasound Image (BUSI), Breast Ultrasound Image (BUI), and Ultrasound Breast Images for Breast Cancer (UBIBC) datasets have been used to build and fine-tune custom and pre-trained CNN models systematically. Custom CNN models have been built, and hence, transfer learning (TL) has been applied to deploy a broad range of pre-trained models, optimised by applying data augmentation techniques and hyperparameter tuning. Models were trained and tested in scenarios involving limited and large datasets to gain insights into their robustness and generality. The obtained results indicated that the custom CNN and VGG19 are the two most suitable architectures for this problem. The experimental results highlight the significance of employing an effective step-by-step approach in image classification tasks to enhance the robustness and generalisation capabilities of CNN-based classifiers.

## 1. Introduction

Artificial Intelligence (AI) is rapidly evolving and integrates computational modelling, high-performance computing, and machine learning with big data records. The advent of Convolutional Neural Networks (CNNs) has significantly enhanced these capabilities, leading to a surge in interest from different research communities, including medical diagnosis [1]. Furthermore, Transfer Learning (TL) has extended CNNs’ applicability to scenarios with limited data, leveraging pre-trained models such as ResNet, UNet, and VGGNet [2]. Although the basic CNN architecture is well-established, there are many variants of it, not least the publicly available pre-trained models that enable TL. In a systematic review of 425 studies [3], TL has been used frequently for image classification. In addition to the existence of diverse pre-trained CNNs, many other refinement techniques are used to enhance the model performance, including the number of convolutional layers and their dimensions, finetuning layers and hyperparameters [4, 5], right optimizer from the many available options (like Adam, RMSprop, Nadam, and Adagrad), regularization and model generalization, overfitting prevention [6]. Refining a backbone architecture (like adding convolutional layers) and tuning its hyperparameters has also been used for optimising a CNN model. Likewise, different types of data augmentation techniques have also been used to improve model robustness [7, 8]. Nanni et al. [7] performed a comparative study of 11 augmentation techniques on four small imaging datasets to reduce overfitting. CNN models were trained after data augmentation, and they were merged into ensemble models to enhance the accuracy. Korzhebin et al. [5] conducted a similar comparative study of four augmentation techniques and five pre-trained CNNs on a small dataset comprising 40 images, only illustrating the model-dependent nature of TL and data augmentation.

Over the years, different alternatives have been presented for medical image classification as an exemplifying application of the CNN architecture[9]. In the pool of miscellaneous alternatives, the adjustment in CNN models to improve the image classification task is confounding several approaches on CNNs for breast cancer detection, such as Pacal et al. [10] Utilised CNNs to classify ultrasound breast images, highlighting the advantages of simple DA and TL, with the vision transformer model emerging as the top performer. Reenadevi et al. [11] employed a deep ResNet-152 model on histopathology images, achieving high accuracy. Similarly, Salama et al. [12] proposed a CNN-based framework for breast cancer segmentation and classification in mammograms, using the MIAS, DDSM [13], and CBIS-DDSM datasets [14]. Al-Dhabyani et al. [15] demonstrated the effectiveness of GAN-based DA in classifying breast cancer from ultrasound images, with NASNet. Saber et al. [16] used TL of pre-trained CNNs for breast cancer detection and classification in mammograms, with VGG16 outperforming existing models after preprocessing and DA. Kumar et al. [17] proposed an approach for ultrasound image classification, achieving higher accuracy with a combination of DA, TL with BreastNet18, and a Cubic SVM. Castro-Tapia et al. [18] explored multi-class breast cancer classification in mammograms using GoogLeNet, achieving superior performance across various metrics. Sharma et al. [19] applied TL with DenseNet for histopathology image classification, showcasing the effectiveness of CNNs and TL. Arooj et al. [4] customised AlexNet using TL and DA for ultrasound and histopathology images, achieving high accuracy. Similarly, Ayana et al. [20] examined TL approaches for ultrasound image diagnosis, emphasising superior performance over non-transfer methods with various preprocessing techniques.

While applying a CNN-based model for image classification, one may overlook some of the many model enhancement techniques or their combinations in an unorganised way, making the effort inefficient. A common framework is a justified need to find the best approach for image classification using deep learning. A common framework can reduce the risk and realise the opportunity by facilitating comparison between methods and results, contributing to quality control, and upscaling. Sijie et al. [21] and Cui et al. [22] have argued for establishing and using a common approach in AI-assisted image classification that can arrange refinement techniques in a step-by-step manner to get the most out of each CNN architecture. Similarly, a common problem encountered by AI learners and researchers is that they stop experimenting with alternative architectures, refinement techniques, and data augmentation techniques after achieving decent results. The existence of diverse evaluation metrics and visualisations often confuses AI researchers too. Hence, it appears a common mistake for AI researchers is to rely on a narrow range of or maybe even just one metric or visualisation for evaluation. Unlike traditional disciplines like physics and economics, AI does not have a clear and standardised framework wherein image-based classification using CNN. Considering these, this study aims to propose a comprehensive approach as a standardised framework for optimising the use of CNNs in image classification.

The approach proposed in this paper encourages image classification tasks to move in this direction by highlighting how a standardised framework aids in selecting the most suitable CNN architecture and refinement techniques for specific tasks. This study emphasises that an approach can enhance image classification tasks by providing a clear framework with the exemplary application of breast cancer detection. In other words, the study contributes to the domain of image classification using CNN in several key ways: i. It highlights the need for a standardised approach to reduce redundancy and foster synergy in CNN-based image classification. ii. It offers a detailed framework for selecting and optimising CNN architectures, including refinement techniques and data augmentation methods. iii. It also provides insights into how such a standardised approach can enhance AI education by clarifying objectives and improving the learning experience for students and educators. This approach could also serve as a roadmap for future advancements in image classification and the fast-growing field of image-based computational applications in healthcare. Furthermore, it may be adapted for other classification tasks, ensuring similar levels of accuracy and methodological rigour. The experimental findings emphasise the importance of selecting appropriate tuning strategies in image classification tasks to improve the robustness and generalisation of CNN-based classifiers.

The rest of the paper is organized as follows: the Materials and Methods used in this study are presented in Section 2; the results of the adopted models are discussed in Section 3; the discussion on the models’ performance is provided in Section 4; and the conclusions of the study are summarised in Section 5.

## 2. Materials and Methods

This study introduces a step-by-step guide for building classification systems with medical images and CNNs, using exemplary breast cancer detection. The study highlights the potential of deep learning DL) and TL in improving the accuracy and efficiency of image classification for cancer detection, proving their vital role in early diagnosis and treatment. The adopted computational experimentation includes building, refining, and experimenting with custom and pre-trained CNN models, incorporating transfer learning (TL), data augmentation (DA), hyperparametric tuning, and ensemble modelling. These models are tested on datasets with varying sizes, clarity, purity, and exposure to seen or unseen data (seen data refers to the dataset(s) used during training, validation, and testing phases where unseen data refers to dataset(s) not used in training, employed to evaluate model performance and generalization on new, previously unencountered images, ensuring robust evaluation metrics and visualisations.

In this study, the three datasets: BUSI, BUI, and UBIBC are used, where UBIBC is a large dataset with high-quality images in PNG format, BUSI is a medium-sized dataset with quality images in PNG format, and BUI is a smaller dataset with slightly blurred images in BMP format. A standardising criterion was applied such that all images were converted to a uniform resolution of 224×224 pixels, standardised to grayscale, and saved in PNG format to ensure consistency. Class imbalances were addressed using the Synthetic Minority Over-sampling Technique (SMOTE), and pixel values were normalised between 0 and 1 to improve model convergence. DA methods were also employed to enhance data diversity and model robustness. While multiple imaging approaches, such as MRI and X-rays, are used for breast cancer detection, this study specifically focuses on ultrasound imaging. Ultrasound is particularly useful for distinguishing between cysts and solid masses in dense breast tissue, making it a preferred choice in specific scenarios. In addition, in this study, several standardised evaluation metrics were used for model performance, including accuracy, precision, recall, F1-score, and AUC. Accuracy measures the overall correctness of classifications, while precision reflects the proportion of true positives among predicted positives. Recall focuses on detecting true positives, and the F1-score balances precision and recall, especially in imbalanced datasets. Similarly, AUC evaluates the model’s ability to differentiate between classes. Visualisations like confusion matrices and ROC curves complement these metrics, ensuring clear and standardised interpretation for both experts and non-experts.

Furthermore, a series of breast cancer image classifiers were built systematically with custom and pre-trained CNN architectures using with and without TL. To sum up, the study implemented a thorough journey of exploring different convolution blocks, frozen layers, normalization techniques, data balancing techniques, DA methods, hyperparameter tuning (activation functions, optimizers, batch sizes, and learning rates), and regularization methods (dropout and early stopping mechanisms), as well as different dataset sizes.

Figure 1 illustrates the steps in the approach to build exemplifying image classifiers. These steps are arranged in the order of building models from scratch with and without TL, DA, hyperparameter tuning, layer tuning, ensembling models, merging datasets with different sizes and degrees of clarity to create fusion datasets, and training models with different amounts of training data, and evaluating their performance for both seen and unseen data. The process begins by developing base models from scratch without utilising TL. These models were first trained with the BUSI dataset without TL, then TL was employed by importing pre-trained models and fine-tuning them. DA techniques were subsequently applied to diversify the training dataset. This step was followed by hyperparameter tuning and layer tuning to optimise the models. Ensemble models were constructed by combining multiple models. Fusion datasets were created by merging datasets from different sources.

**Figure 1:**
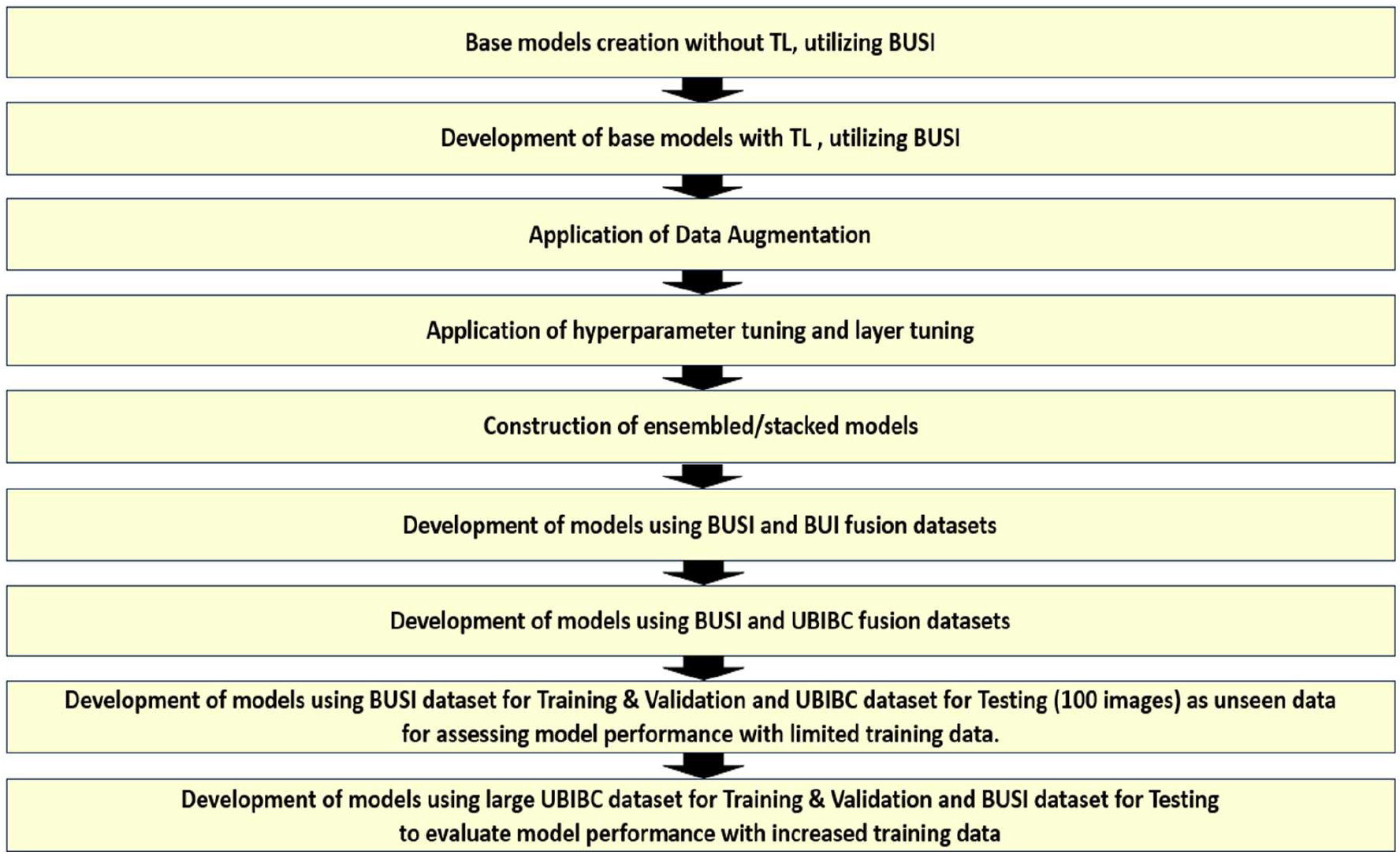
Conceptual workflow of the framework.

In this work, the combination of BUSI and BUI datasets and BUSI and UBIBC datasets was used, where models were first tested using the same dataset by splitting into training, validation, and testing data. This allowed performance evaluation of the model from the same dataset. Following this, the models were tested using unseen data and evaluated for how well the models were generalised to new, previously unencountered data. In the next step, models were tested on a larger training dataset and hence the evaluation of their performance. With these steps, the presented work is structured as a framework which provides a systematic step-by-step approach to CNN-based image classifier development and evaluation, ensuring a range of alternative techniques with the available datasets in an image classification task to achieve optimal performance in a task-specific and context-specific manner.

### 2.1 Data Preparation

Different datasets with different numbers of image data are used in this study, as shown in Table 1. TO illustrate to images that have been used in this study, figures 2 to 4 show the random representative images of different datasets. To build the CNN models, each dataset was systematically partitioned into training, validation, and testing subsets to prevent data overlap across these stages. Additionally, in some experiments, distinct datasets were used for training, validation, and testing to assess model performance based on unseen data (data belonging to different datasets), mitigating overfitting and enhancing overall performance.

**Table 1:** Different datasets used in the research study.

| Dataset | Images |  | Class |  |  |
| --- | --- | --- | --- | --- | --- |
|  | Total | Format | Benign | Malignant | Normal |
| BUSI - Breast UltraSound Images | 780 | PNG | 437 | 210 | 133 |
| BUI - Breast Ultrasound Images | 250 | BMP | 100 | 150 | - |
| UBIBC - Ultrasound Breast Images for Breast Cancer | 9016 | PNG | 4574 | 4442 | - |

**Figure 2:**
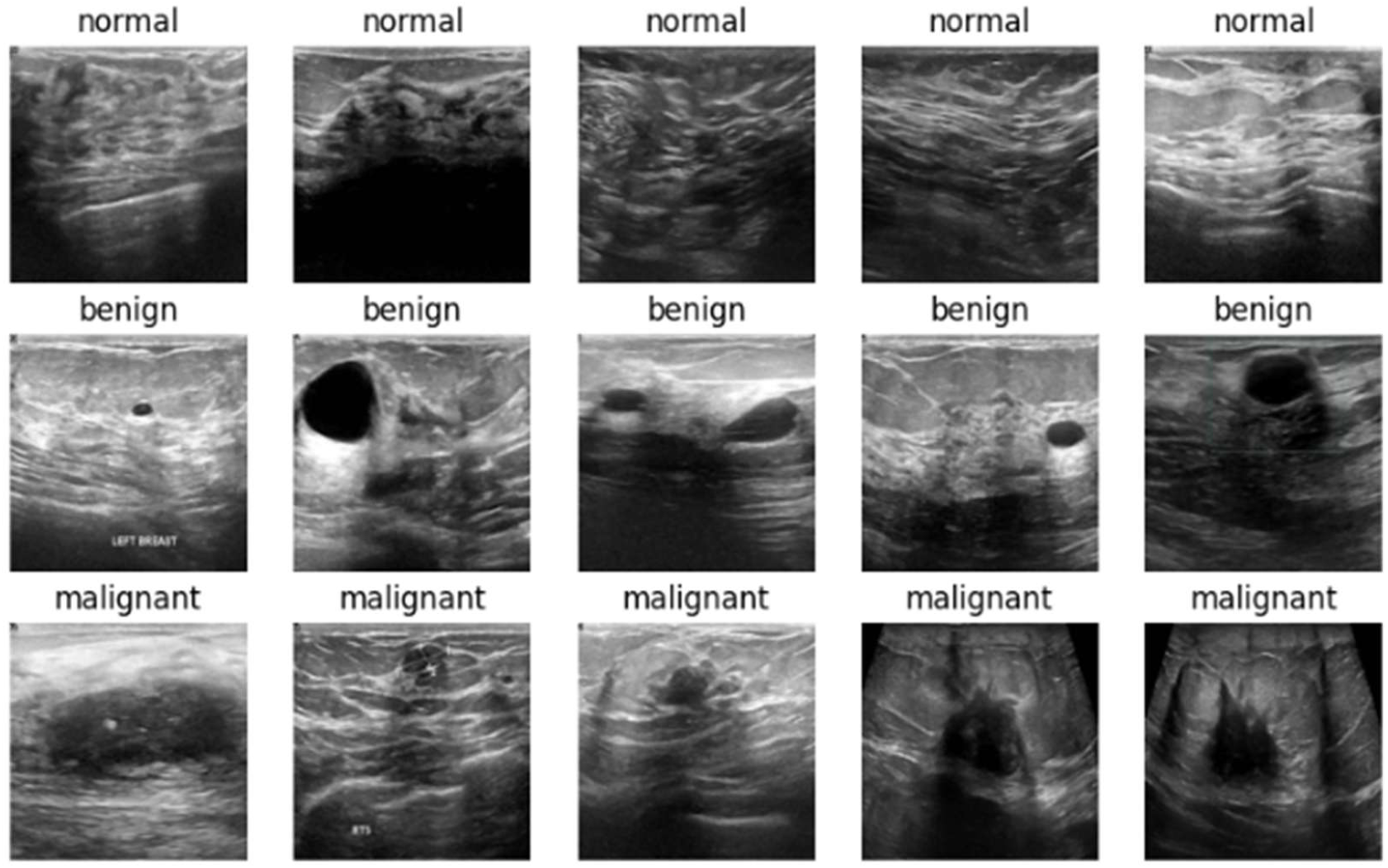
Sample images of BUSI - Breast Ultrasound Images.

**Figure 3:**
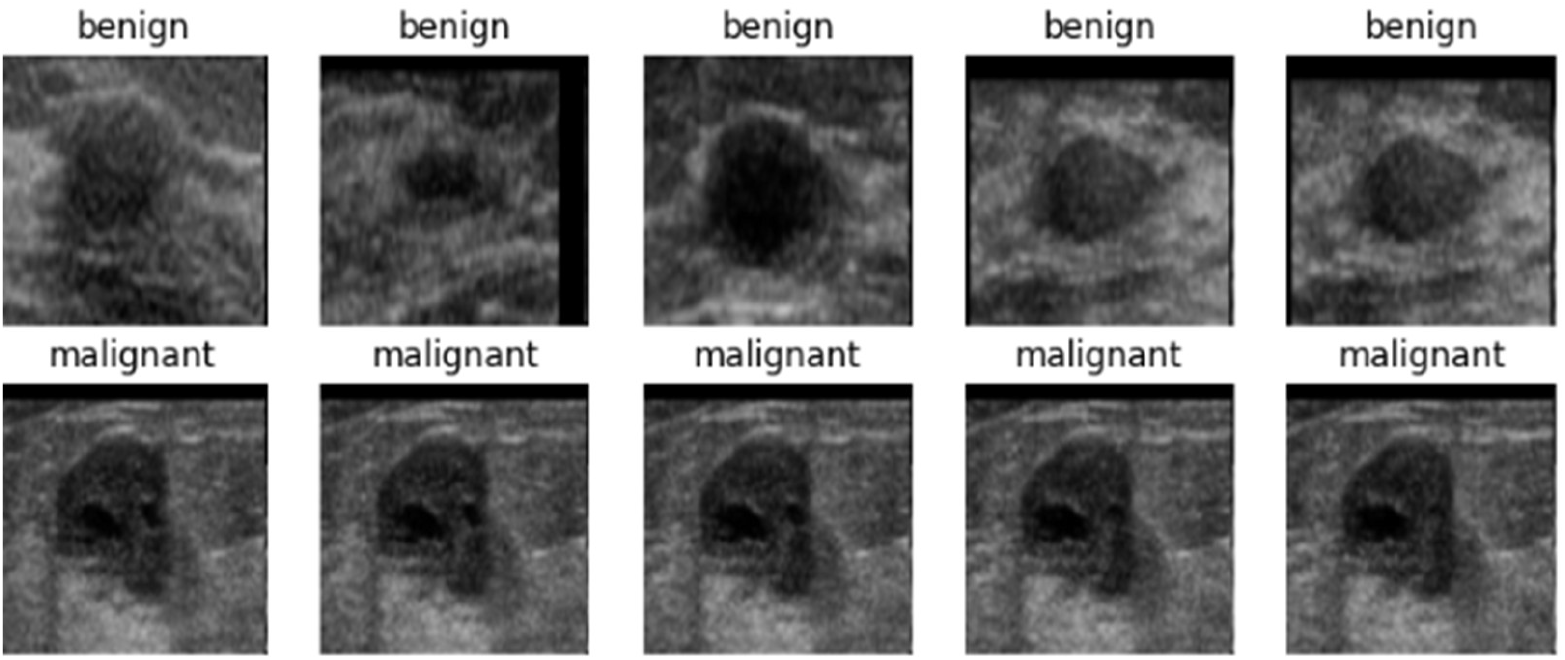
Sample images of BUI - Breast Ultrasound Images.

**Figure 4:**
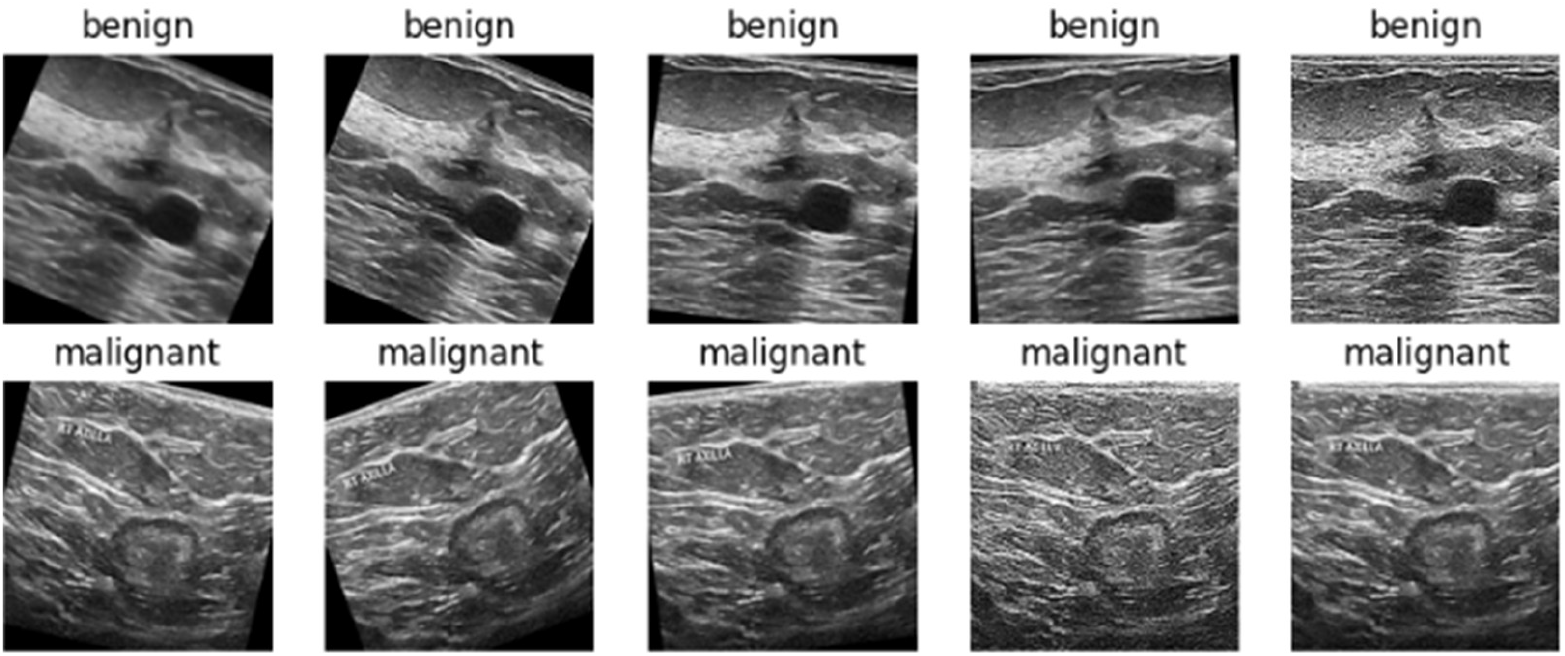
Sample images of UBIBC - Ultrasound Breast Images for Breast Cancer.

### 2.2 Data Preprocessing

Data preprocessing has been applied to prepare the dataset for the models. As a part of data preprocessing, image formats are standardised by converting all images to grayscale, resizing the images into 224×224 pixels, and saving them in PNG (Portable Network Graphics) format. To address class imbalances, SMOTE have been applied. Besides, Pixel values are normalised to a range between 0 and 1 to aid in model convergence. Additionally, DA techniques, including random cropping, flipping, shifting, shearing, rotation, and zooming, have been used to increase the diversity of the training data.

### 2.3 Model Development

Out of the many CNN architectures available in the public domain, a shortlist was devised for breast cancer classification based on some of the recommendations in the scholarly articles [3, 6, 10, 11, 15, 23]. Custom and pre-trained CNN architectures with and without TL were used to build a series of models, which were enhanced methodically with architectural variations (as shown in Figure 5 and Figure 6), normalisation techniques, and hyperparameter tuning techniques. The selected pre-trained CNN models were initialised with ImageNet pre-trained weights, and their dense custom layers were initialised using GlorotUniform (Xavier uniform) to optimise training efficiency and mitigate gradient-related issues. Various optimisers like SGD, Nadam, and Adagrad were used with the categorical cross-entropy loss as the objective function [6]. Class weights were incorporated using the ‘balanced’ option to address imbalanced datasets. The resulting weights were stored in a dictionary in a class-dependent manner, ensuring that minority classes would be oversampled during model training.

**Figure 5:**
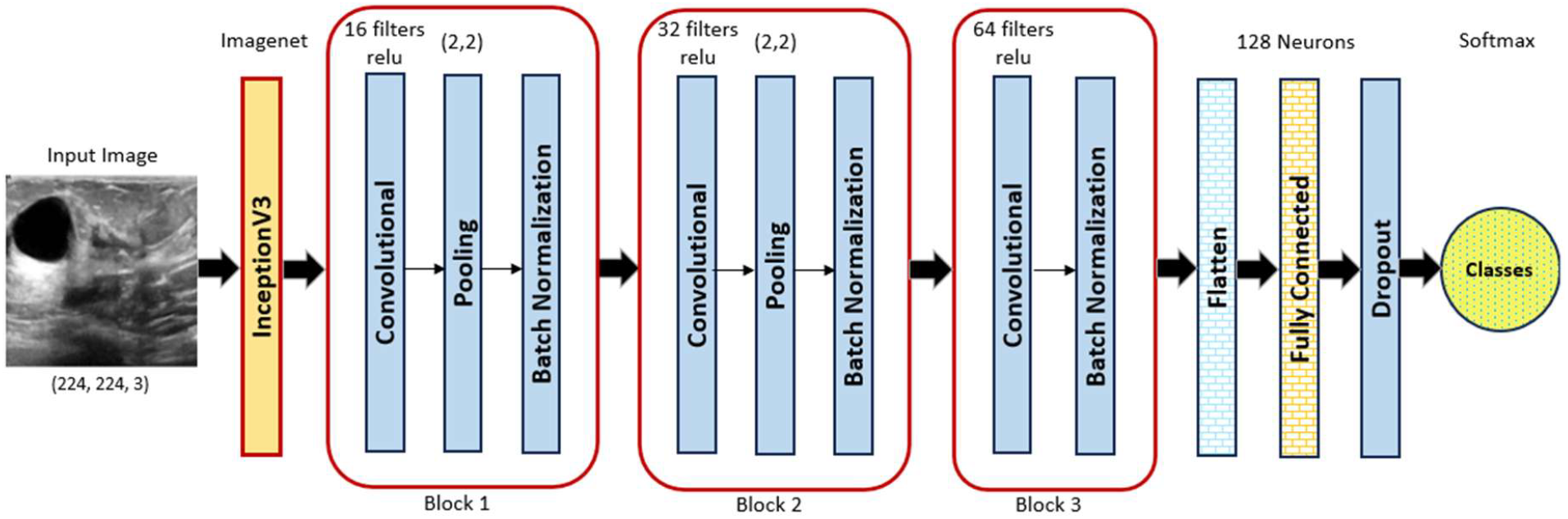
Architecture of Keras Sequential Model with Transfer Learning.

**Figure 6:**
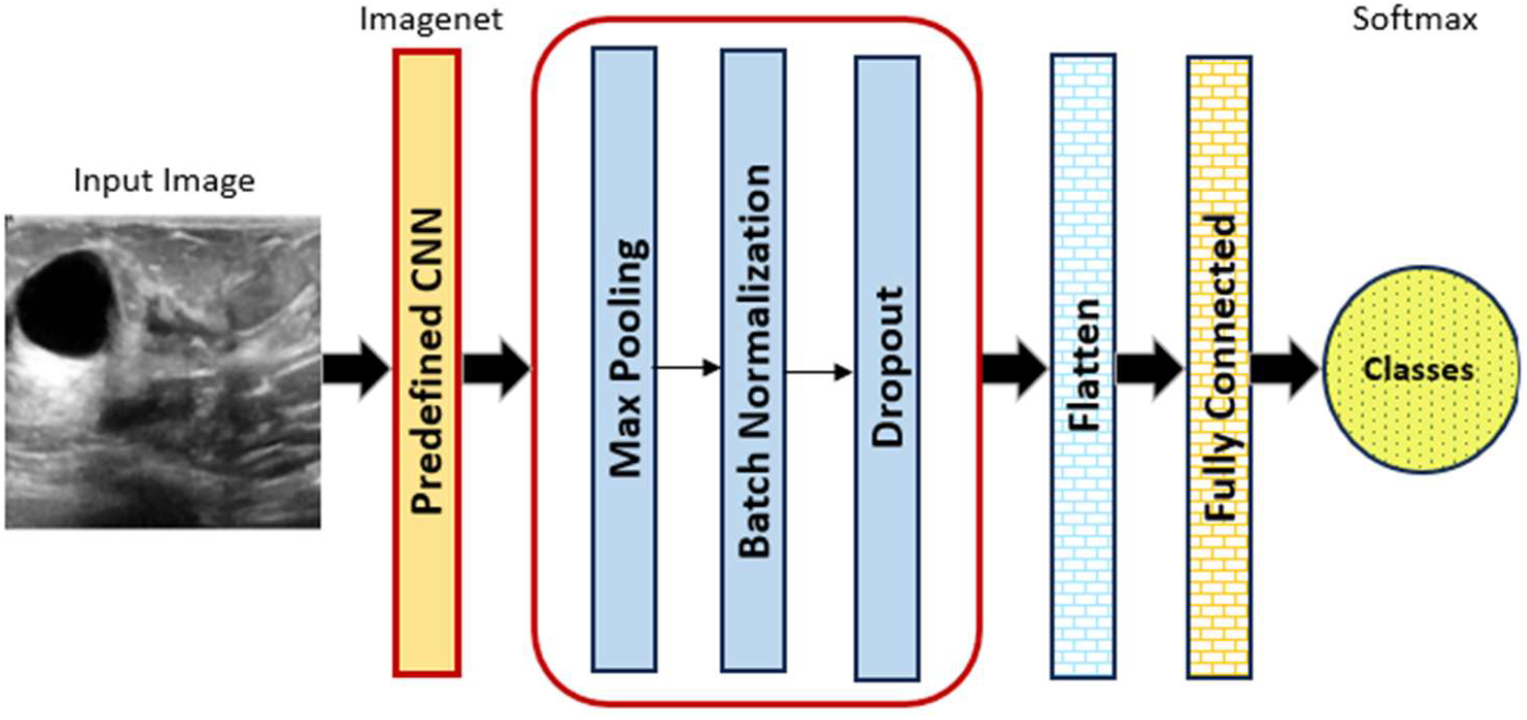
Architecture of Predefined CNN Models with Transfer Learning.

Classification models were trained using different strategies involving distinct datasets, including hybrid datasets created by fusion. At first, the BUSI and BUI datasets were fused before being meticulously split into training (70%), validation (15%), and test (15%) subsets [10]. A similar approach was applied to the fusion of the BUSI and UBIBC datasets. In one qualitatively different case not involving hybrid datasets, the BUSI dataset was split into two subsets for training and validation, while selected UBIBC images were used for testing. Similarly, models were trained and validated on a large dataset (UBIBC) and evaluated against smaller datasets (BUSI). To prevent overfitting, early stopping was used during model training by monitoring each model’s validation loss and accuracy to gain insights into its learning dynamics and terminating its training when it was observed to diverge from its training loss. A Learning Rate scheduler was utilised to adjust (decrease) the learning rate by a factor of 0.5 if the validation loss stagnated for 5 consecutive epochs. Each trained model was tested on a test dataset, and in terms of a spectrum of key metrics, including testing accuracy, precision, recall, F1-score, and AUC-ROC Curve [6]. Confusion matrices were also built to supplement these individual metrics by providing a holistic assessment of the model’s performance. Figure 5 shows the Sequential model architecture comprising a pre-trained InceptionV3 base model followed by additional layers for feature extraction and classification.

The additional layers begin with two convolutional layers with 16 and 32 filters, respectively. Their outputs pass through ReLU activation functions, average pooling layers, and batch normalisation functions. The third additional layer is a convolutional layer with 64 filters, followed by a flattening layer and a dense layer comprising 128 units with ReLU activation and dropout regularisation. The final layer simply comprises two units with a softmax activation for binary classification. In addition to dropout regularisation, some of the dense layers are regularised through L2 regularisation with a coefficient of 0.01. Figure 6 shows the architecture of a general pre-defined CNN. In each specific implementation of this general architecture, the model’s pre-trained weights were derived from the ImageNet dataset and finetuned by freezing different layers, for example, freezing the 10 layers before the last layer.

## 3. Experimental Analysis

Following the conceptual framework described in previous sections, a series of machine-learning experiments were conducted in alignment with the presented method to build, optimise, and evaluate the exemplary application of breast cancer image classifiers. These experiments, with different alternatives, were conducted for image classification tasks, moving in this direction by the importance of how a standardised step-by-step approach helps in selecting the most suitable CNN architecture and enhancement techniques for domain-specific tasks. For the experimental analysis to support the step-by-step approach of CNN model development, three datasets, along with combination data augmentation techniques, were used for model training.

### 3.1 Experiment 1: Base models with and without Transfer Learning

At first, 10 base models were created without TL, encompassing architectures such as Sequential in Keras (custom) and pre-defined models (Shortlisted based on recommendations in the articles [3, 6, 10, 11, 15, 23]. Simultaneously, another 10 base models were developed based on the same architectures with TL, with the weights being initialised with the ‘imagenet’ setting for improved feature extraction as listed in Table 2. For the analysis, the first 10 models without TL performed sub-optimally compared to their counterparts with TL. The base models, without TL, were not learning effectively as the loss and accuracy values did not improve over epochs. On the other hand, the models with TL improved consistently in terms of training loss, indicative of effective learning and generalisation to unseen data. This shows that leveraging pre-trained weights helped the second set of 10 models learn from the training dataset effectively, aligning with expectations. Sequential, EfficientNetB3, VGG19, ResNet152, InceptionV3, and InceptionResNet2 models with TL were notably performant in this experiment.

**Table 2:**
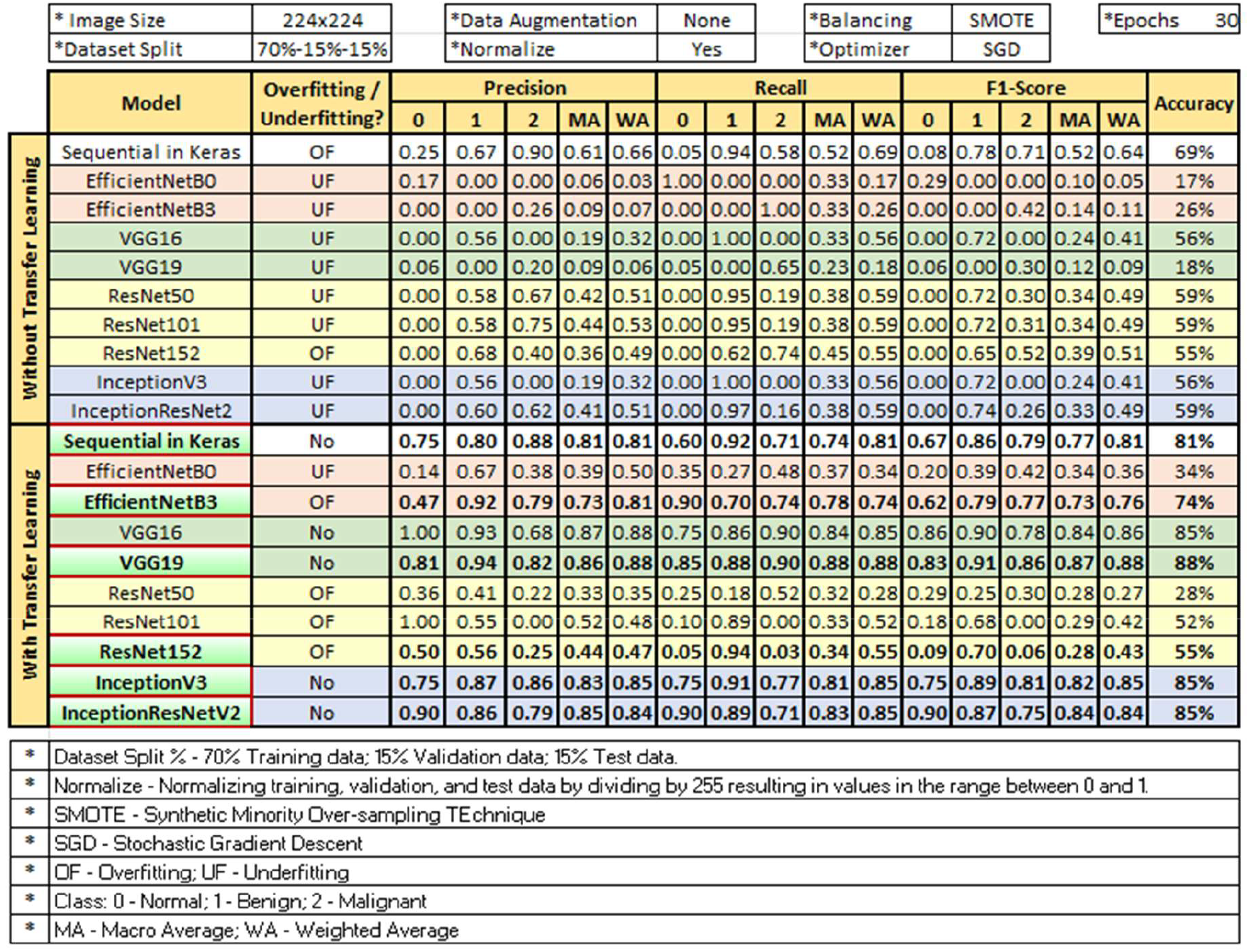
Base models with and without Transfer Learning.

The evaluation metrics: precision, recall, F1-Score, and accuracy were analysed for each model for each class (0: ‘normal’, 1: ‘benign’, 2:’malignant’) as listed in Table 2. The table shows that TL has built efficient classifiers, particularly evident in the precision, recall, and F1-score metrics. For instance, the VGG19, InceptionV3, InceptionResNetV2, and Sequential in Keras models with TL achieved levels of accuracy of 88%, 85%, 85%, and 81%, respectively.

### 3.2 Experiment 2: Application of data augmentation techniques

In the next stage, various DA techniques were utilised, where DA combinations were shortlisted after reviewing different articles [2, 4, 5, 8, 10-12, 16-18, 23, 24], summarised in Table 3. Horizontal and vertical flips, rotation, shear, zoom, width and height shift and cropping were shortlisted due to their efficacy in the reviewed articles. Four combinations of them were applied to Sequential in Keras, EfficientNetB3, VGG19, ResNet152, InceptionV3, and InceptionResNet2, which were the top performers in the first experiment. Among these combinations, horizontal and vertical flip, horizontal and vertical shift, shear, rotation, zoom, and cropping achieved high levels of accuracy as summarised in Table 4.

**Table 3:**
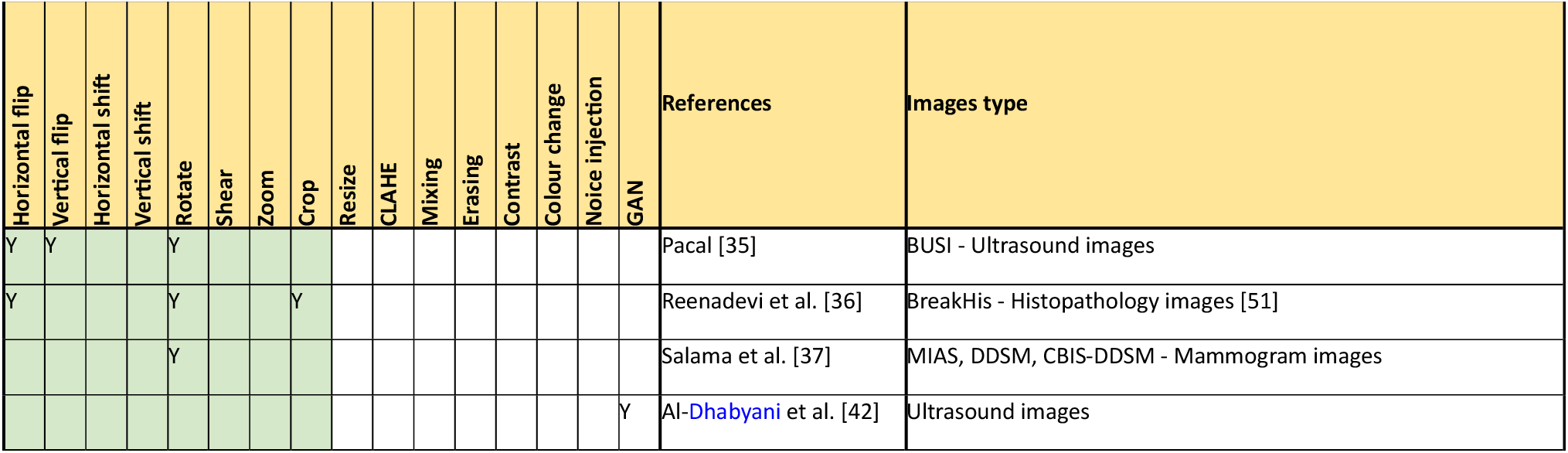
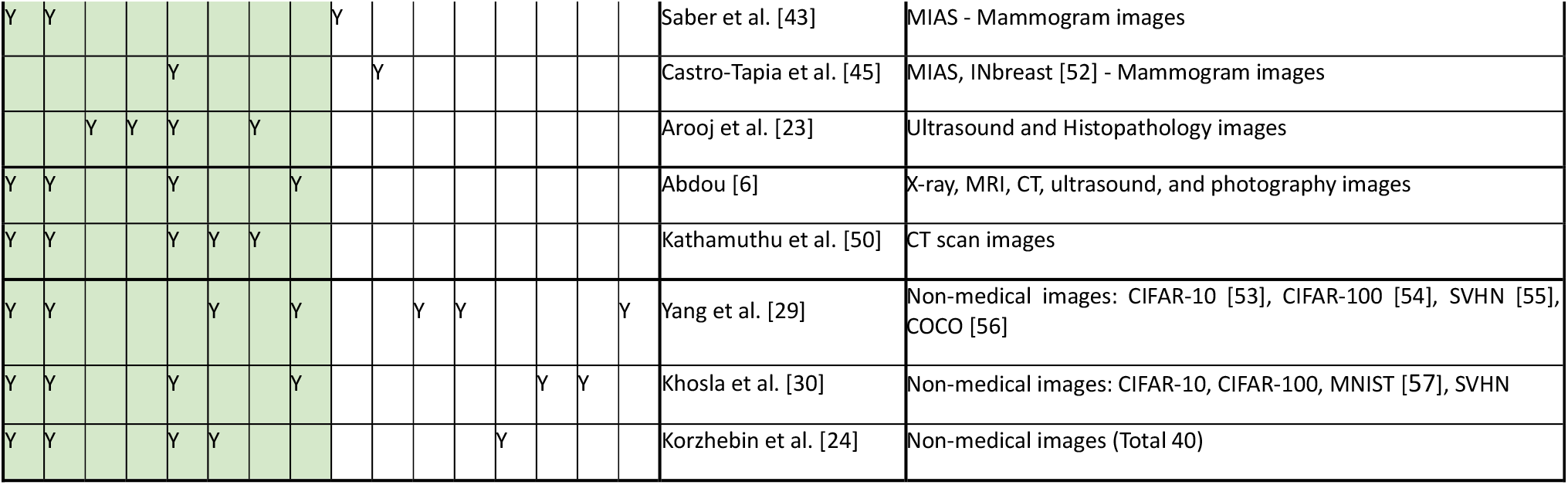
Data Augmentation Techniques.

**Table 4:**
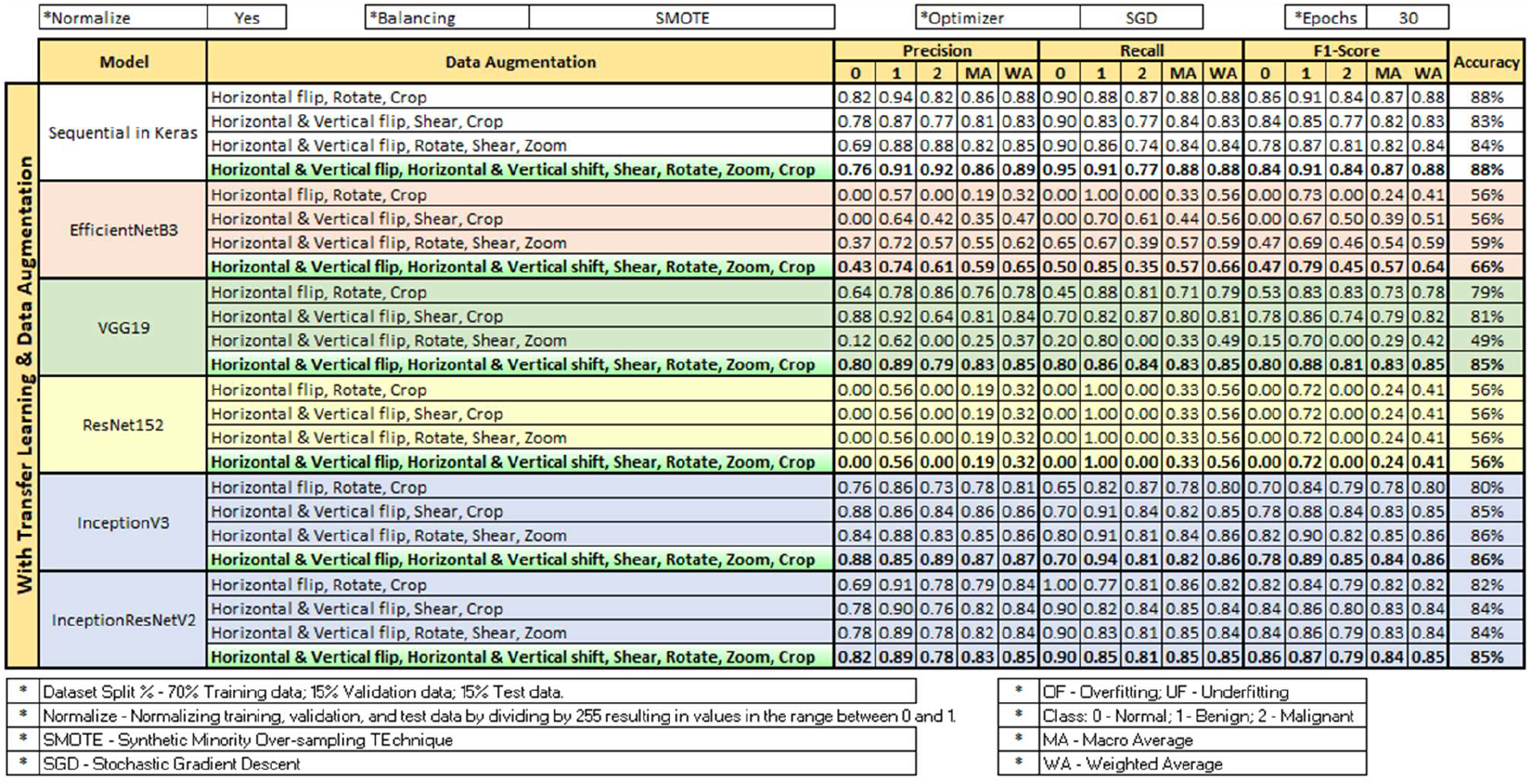
Models with Transfer Learning, Data Augmentation, and one dataset (BUSI)

### 3.3 Experiment 3: Bayesian hyperparameter tuning

Bayesian optimisation enabled hyperparameter tuning affecting the models systematically, tailored by the findings of Balaha et al. [6]. The experiment identified contributing factors to model robustness are two activation functions (relu and softmax), L2 regularization with a coefficient of 0.01, batch normalization (16,32), early stopping, dropout (0.5), convolution blocks (16, 32, 64 filters) and use of dynamic learning rate scheduler to reduce the default learning rate (0.001) by a factor of 0.5. Notably, different TL-based models worked well with different optimisers (AdaGrad, SGD, Nadam), emphasising the nuanced impact of hyperparameter tuning on different architectures. Different patterns of frozen layers with selective freezing, such as freezing the last 10 layers and freezing the penultimate layer only were experimented with [3, 5]. However, this did not improve the results; in some cases, it was a counter-productive effect. The analysis, presented in Table 5, supplemented by Figure 7, shows that Nadam is an effective optimiser for Sequential in Keras, InceptionV3, and InceptionRes-NetV2 models, while Adagrad performed well for VGG19, EfficientNetB3, and ResNet152.

**Table 5:**
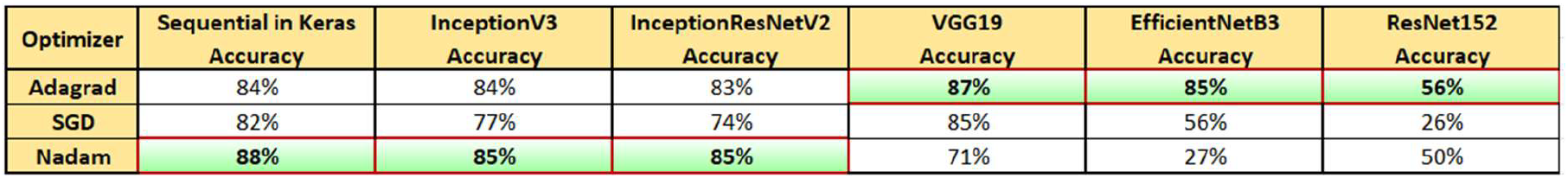
Models with Transfer Learning, Data Augmentation, Tuning, and one dataset (BUSI)

**Figure 7:**
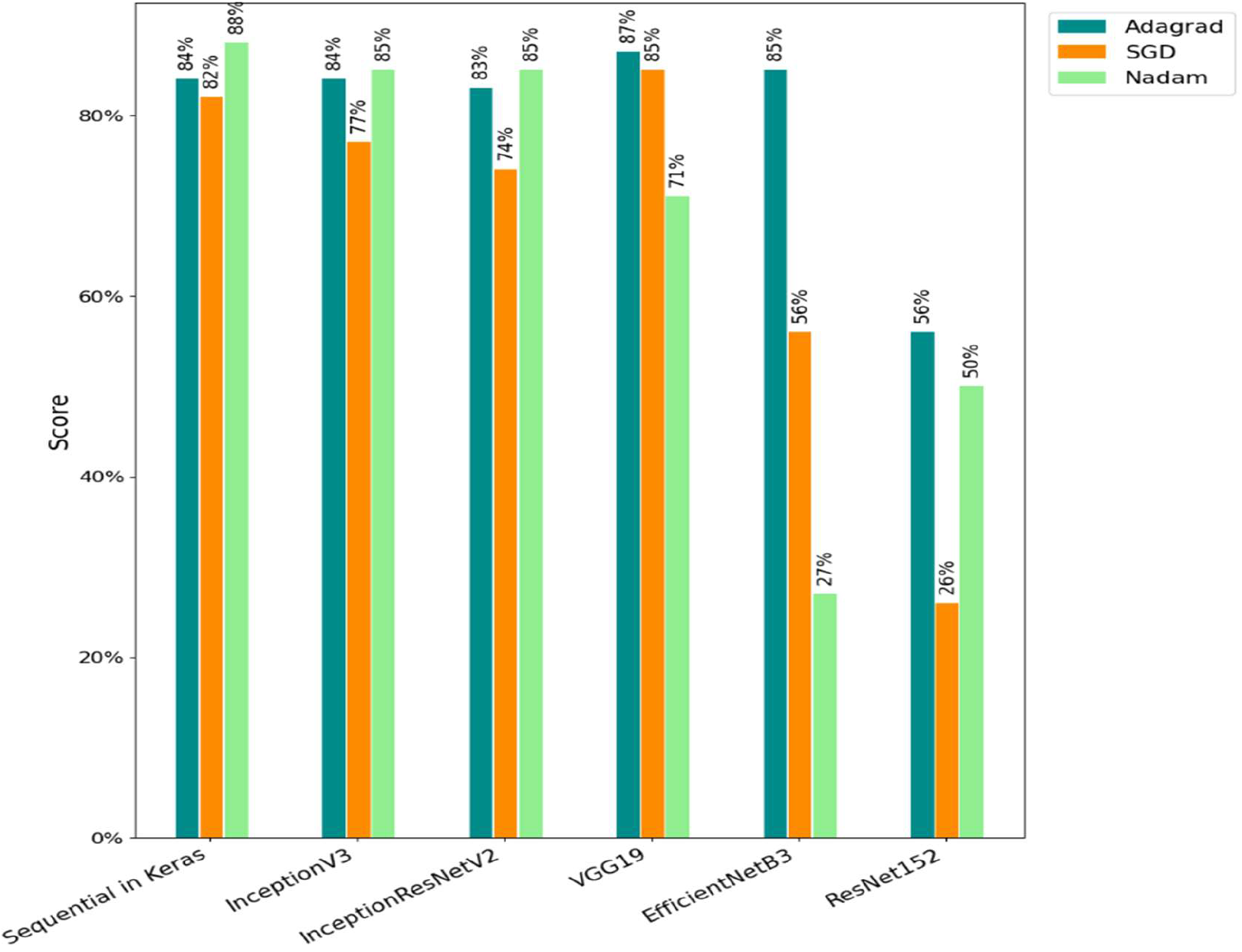
Optimiser selection based on the accuracy of models.

### 3.4 Experiment 4: Stacked (ensemble) models

In the next step, the effectiveness of stacked models for image classification was analysed. Initial attempts involved stacking VGG19, used for feature extraction, on Sequential in Keras, but suboptimal results were observed. Subsequently, a more complex model was devised by stacking VGG19 and InceptionResNetV2, which concatenated their extracted features before feeding them to a final series of fully connected layers for classification. However, this sophisticated approach also did not achieve significantly better results. The steps highlighted that there have been potential difficulties in exploiting an ensemble approach with a relatively small dataset.

### 3.5 Experiment 5: Splitting BUSI/BUI fusion dataset into training, validation, and testing subsets

Furthermore, in the next step, a comprehensive approach was implemented to enhance the performance of DL models by combining BUSI and BUI datasets and splitting the resulting BUSI/BUI fusion with diverse datasets into training (70%), validation (15%), and test (15%) subsets. The analysis has shown that models based on the VGG19 and InceptionResNetV2 architectures were finetuned by TL and empowered by DA, hyperparameter tuning, and class balancing with class weights achieved a slightly higher level of accuracy at the cost of a diminished AUC score. Additionally, applying GlorotUniform (also known as Xavier uniform) weight initialisation for the dense custom layers yielded a marginal accuracy boost in some models at the cost of a diminished AUC score.

### 3.6 Experiment 6: Fusion of BUSI and UBIBC datasets

#### 3.6.1 Experiment 6(a): Splitting fusion dataset into training, validation, and testing subsets

A combined dataset created by the fusion of BUSI and UBIBC datasets was meticulously split into training (70%), validation (15%), and test (15%) subsets. Models constructed using Sequential in Keras, VGG19, InceptionV3, and InceptionResNetV2 exhibited better testing results in terms of precision, Recall, F1-score, AUC, and Accuracy. Notably, Sequential in Keras and VGG19 emerged as the top performers in this experiment. The results of this experiment are listed in Table 6.

**Table 6:** Models with Transfer Learning, Data Augmentation, Tuning, and two datasets (BUSI & UBIBC)

|  |  |  |  |  |  |  |  |  |  |  |
| --- | --- | --- | --- | --- | --- | --- | --- | --- | --- | --- |
| Combined BUSI (647 images) and UBIBC (446 original images) datasets, and split the total 1093 images into training (70%), validation (15%) and test (15%) datasets. |  |  |  |  |  |  |  |  |  |  |
| Model | Optimizer | Precision |  | Recall |  | F1-Score |  | AUC |  | Accuracy |
|  |  | MA | WA | MA | WA | MA | WA | 0 | 1 |  |
| Sequential in Keras | Nadam | 0.95 | 0.96 | 0.96 | 0.96 | 0.95 | 0.96 | 0.99 | 0.99 | 96% |
| VGG19 | Adagrad | 0.93 | 0.94 | 0.94 | 0.94 | 0.93 | 0.94 | 0.99 | 0.99 | 94% |
| InceptionV3 | Nadam | 0.89 | 0.91 | 0.91 | 0.90 | 0.89 | 0.90 | 0.98 | 0.98 | 90% |
| InceptionResNetV2 | Nadam | 0.91 | 0.88 | 0.80 | 0.85 | 0.82 | 0.84 | 0.99 | 0.99 | 85% |

#### 3.6.2 Experiment 6(b): Using different datasets for training/validation and testing

In this setup, the BUSI dataset was used for Training & Validation, while UBIBC was only used for Testing. The models demonstrated robustness by generalising to the unseen dataset (UBIBC) without difficulty. VGG19 achieved an accuracy of 82%, AUC scores of 93% for Benign and Malignant classes, and Precision, Recall, and F1-Score of 84%, 82%, and 82%, respectively, as listed in Table 7.

**Table 7:** Models with Transfer Learning, Data Augmentation, Tuning, two datasets (BUSI & UBIBC)

|  |  |  |  |  |  |  |  |  |  |  |
| --- | --- | --- | --- | --- | --- | --- | --- | --- | --- | --- |
| BUSI dataset for Training & Validation (647 images); UBIBC dataset for Testing (100 images) |  |  |  |  |  |  |  |  |  |  |
| Model | Optimizer | Precision |  | Recall |  | F1-Score |  | AUC |  | Accuracy |
|  |  | MA | WA | MA | WA | MA | WA | 0 | 1 |  |
| Sequential in Keras | Nadam | 0.81 | 0.81 | 0.79 | 0.79 | 0.79 | 0.79 | 0.83 | 0.83 | 79% |
| VGG19 | Adagrad | 0.84 | 0.84 | 0.82 | 0.82 | 0.82 | 0.82 | 0.93 | 0.93 | 82% |
| InceptionV3 | Nadam | 0.81 | 0.81 | 0.81 | 0.81 | 0.81 | 0.81 | 0.88 | 0.88 | 81% |
| InceptionResNetV2 | Nadam | 0.81 | 0.81 | 0.81 | 0.81 | 0.81 | 0.81 | 0.91 | 0.91 | 81% |

Analysing both the fusion, it has been observed that the hybrid dataset allowed the models to generalise and perform well on the combined test dataset. The higher accuracy and evaluation metrics, as presented in Figure 8, support the notion that merging two distinct datasets into a hybrid dataset for all stages is superior to using them separately at different stages.

**Figure 8:**
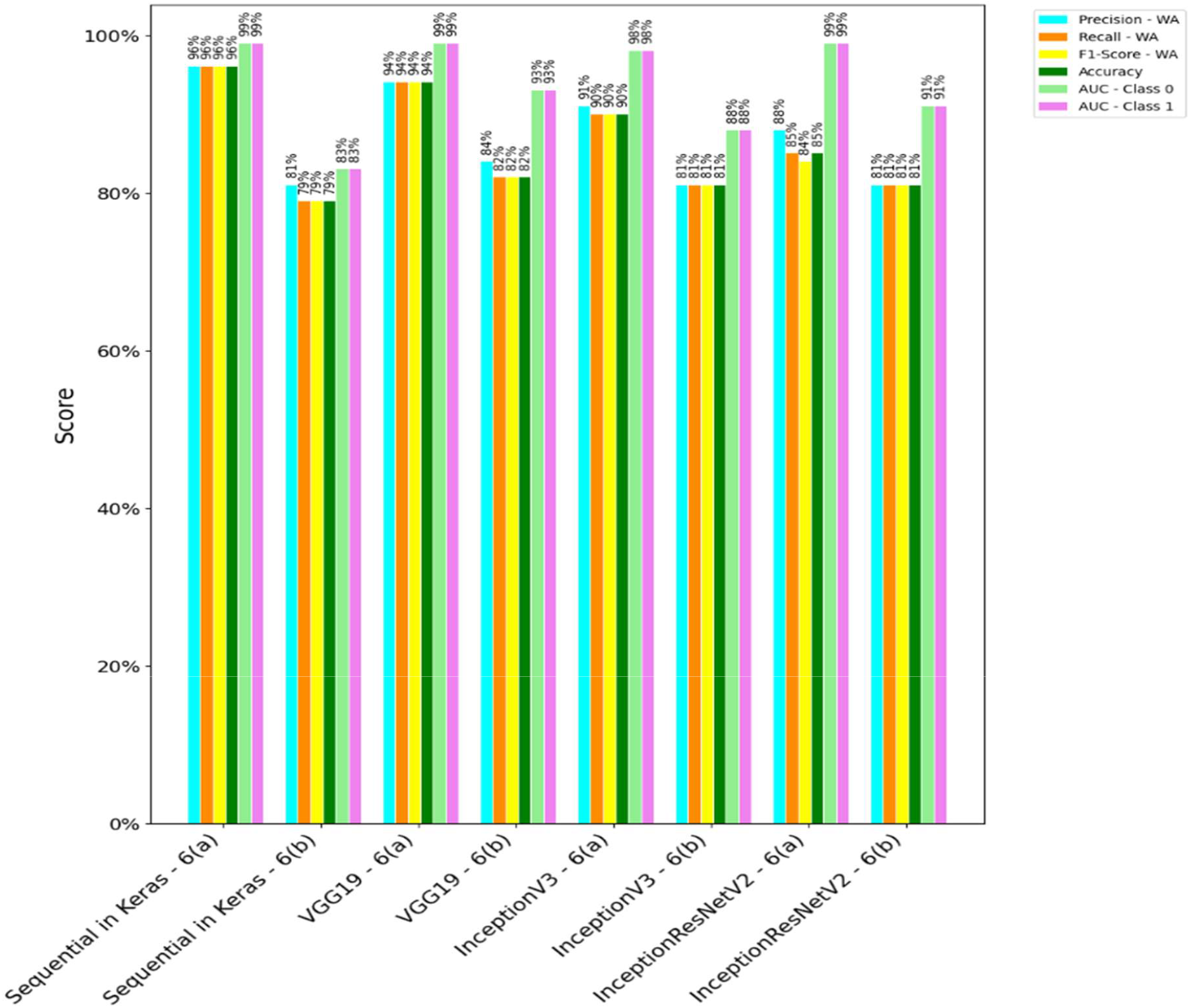
Comparing the Metrics of Experiments 6(a) and 6(b).

### 3.7 Experiment 7: Models using a large dataset (UBIBC) for training & validation, and a small dataset (BUSI) for testing

This experiment involved training/validating models on a large dataset from UBIBC and testing their performance against smaller datasets from BUSI). The models based on the Sequential and VGG19 architectures fine-tuned by TL demonstrated better performance with metrics Precision (0.93), Recall (0.92), F1-Score (0.92) and Accuracy (92%), underscoring their ability to make positive predictions and effectively capture positive instances. AUC values of 0.99 reflect excellent discriminatory power, particularly in Benign and Malignant cases, as shown in Figure 9.

**Figure 9:**
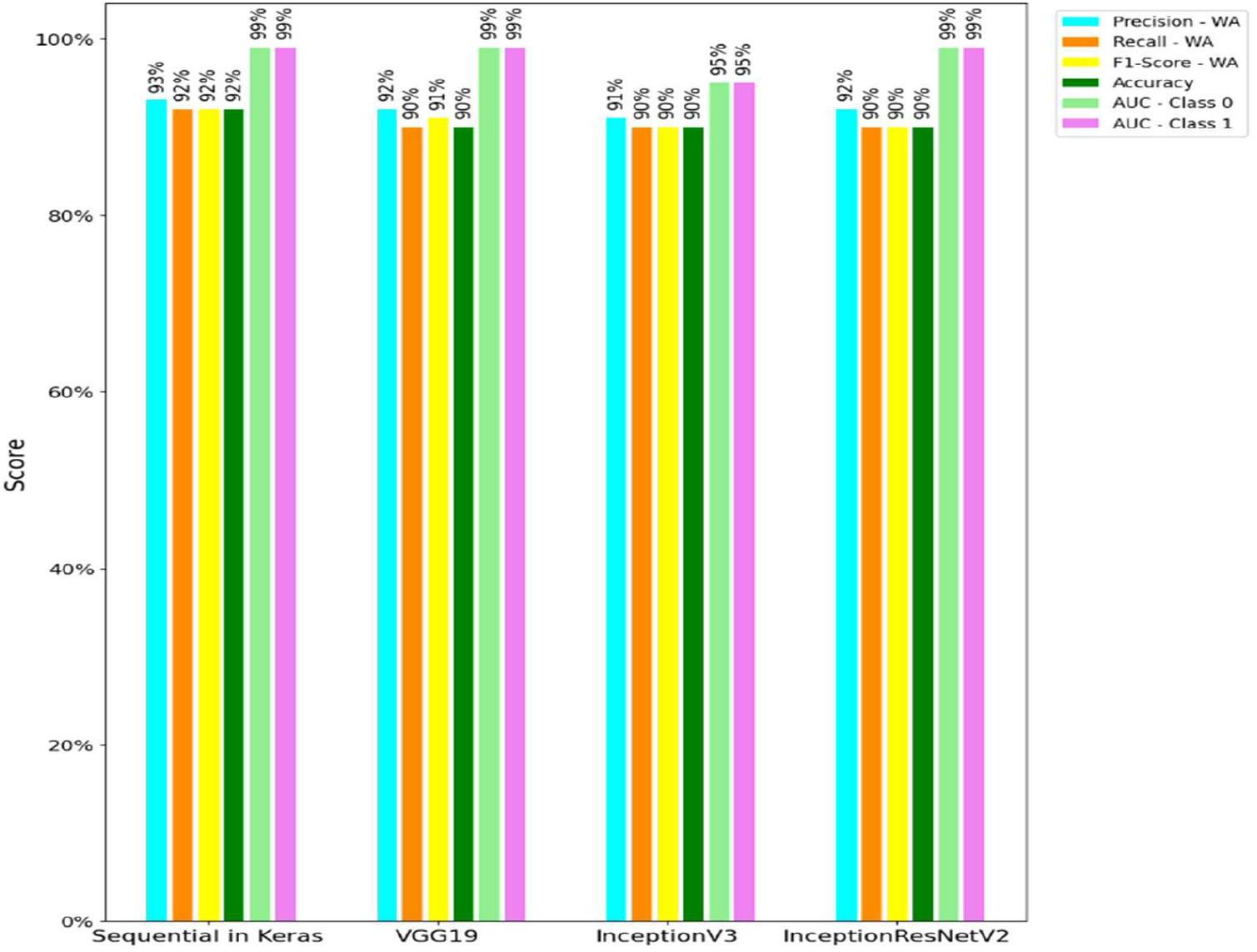
Metrics for UBIBC (Training & Validation) and BUSI (Testing).

## 5. Experimental Result Arguments

This study extends the understanding of stepwise implementation techniques for image classification and their significant impact on model performance. Different alternative experiments were implemented and analysed to explore an efficient way of developing an image classifier. The results of experiment 1 highlighted that models with TL generally outperform those without TL, demonstrating the effectiveness of leveraging pre-trained weights from large datasets like ImageNet. This finding has also been aligned with the existing literature [3-6, 10-12, 15-17, 19, 20, 23, 25], reinforcing the value of TL in enhancing feature extraction and generalisation, especially in medical imaging, where a large volume of data is not often available. The results of experiment 2 further emphasised the importance of DA techniques in improving model robustness and performance. But, there is the limitation that some combinations of DA techniques are better than others, so applying them blindly without considering the nuances of a particular problem/dataset may generate lower accuracy models. However, the combination of horizontal and vertical flips, shifts, shear, rotation, zoom, and cropping proved particularly effective in this study. These techniques have been validated in other studies [3-6, 10-12, 15-17, 19, 20, 23, 25] as well, showing their efficacy in creating diverse training samples that enhance model generalisation.

Furthermore, the exploration of hyperparameter tuning has also been implemented in experiment 3. The results were indicative of the nuanced effects of different optimisers (AdaGrad, SGD, Nadam) and configurations, such as learning rates, dropout rates, batch sizes, convolution blocks, batch normalisation, early stopping, L2 regularisation, and activation functions on model performance. This result underscored the necessity of tailored hyperparameter tuning for achieving optimal performance in DL models. For example, different CNN architectures may require different optimisers [6, 18] for optimal performance, and they may require different numbers of frozen layers during finetuning [3, 5]. Going further, stacked ensemble models from Experiment 4 did not yield significant performance improvements, highlighting the challenges of working with relatively small datasets. This finding is aligned with the broader understanding that ensemble methods often require large amounts of data to realise their full potential [26]. The experiment underscored the importance of dataset size and diversity in achieving substantial benefits from ensemble techniques.

Experiments 5 and 6 have focused on the impact of dataset fusion and image clarity on model performance. The improvements observed in models using combined datasets and the superior performance of models on the clearer dataset underscore the critical role of data quality and diversity, suggesting that mixing high-quality images could be a useful procedure in general. The results of experiment 5 highlighted the fact that optimising DL models sometimes involves trade-offs. In that experiment, we managed to achieve a higher accuracy at the cost of a lower AUC score. Experiment 6, particularly the comparison between using combined datasets for training, validation, and testing, and using two distinct datasets for training/validation and testing, produced results that highlighted the importance of exposing a CNN model to diverse examples with distinct characteristics during training for effective generalisation. Experiment 7 demonstrated the robustness of models trained on a large dataset and tested with different smaller datasets. This experiment underscores the potential of large, high-quality datasets in training robust models capable of generalising well to different datasets.

Following the step-by-step approach not only standardises the image classification tasks but also achieves better results than comparable studies, such as the work presented by Balaha et al. [6] which employed the fusion of BUSI and BUI datasets to assess diverse DL architectures: InceptionResNetV2, ResNet152, VGG19 and Xception, each utilising a specific optimiser. Their study revealed distinct performance outcomes, with InceptionResNetV2 achieving the highest accuracy (88.45%), Precision (89.80%), Recall (87.04%), F1-score (88.38%), and AUC at 97.04%. In contrast, this research also highlighted the sequential model’s superiority over other models, as the Sequential model achieved Accuracy (92%), Precision (93%), Recall (92%), F1-Score (92%), and AUC (99%), demonstrating superior accuracy. This model had robust classification, minimised false positives, and shorter training time, emphasising its suitability for medical classification tasks. While the model demonstrates strong performance across multiple datasets, several limitations were also observed, particularly in terms of dataset selection and generalisability. The model’s performance varied when trained on one dataset and tested on another, highlighting the challenge of dataset generalisation. This suggests that while the model performs well on the specific datasets used in this study, its effectiveness may decrease when applied to different datasets without further fine-tuning. This limitation could impact the generalisability of the findings to other applications or imaging modalities, such as mammography or MRI, where data characteristics differ significantly from ultrasound.

## 4. Discussion and Conclusions

The use of CNNs is increasingly common, and this deep-learning architecture is an essential component of image classification. Major challenges in building CNN-based image classification applications are the availability of wider architectures, hyperparameter tuning targets and techniques, and data augmentation techniques. A common approach can help to minimise redundancy, facilitate model comparison, streamline communication, encourage quality control, and promote upscaling. This paper proposes a step-by-step approach for building image classifiers and exemplifies it by reporting how to build CNN models to classify ultrasound breast cancer images within the framework. It underscores the critical roles of TL, DA, hyperparameter tuning, and dataset fusion in achieving robust and high-performing models. The Step-by-Step approach is a checklist to execute combinations relevant to the CNN-based applications with the image classification task. Besides, the wider range of experimentation, analyses and discussions contributes to the broader understanding of DL applications in imaging and offers valuable guidance for future research and applications. Through an extensive exploration of DL architectures, TL, and DA techniques, it has been concluded that incorporating a diverse range of training datasets is crucial for achieving optimal model performance, specifically, using varied datasets, including those with different image qualities and characteristics, significantly enhanced the model’s ability to generalise and perform accurately across various scenarios. The systematic methodology, encompassing TL, DA, hyperparameter tuning, ensemble models, and data diversity, provides both a methodology for and valuable insights into the common challenge of optimising DL models for image classification.

This approach can be a roadmap for future endeavours in improving the fast-growing field of image-based computational medicine. More generally, we hope that the approach will contribute to a standardised development of image classification applications that align with Jerome Bruner’s theory of scaffolded learning [27] which states that when a learner is exposed to medical image classification for the first time, they need active support. By navigating them with the aid of a structured approach, anyone building a CNN-based image classifier gains confidence and independence by following Kolb’s model of experiential learning [28]. For example, if a student is new to image classification, the presented approach would help them retain and recall aspects of their first experience of building and training a CNN. While reflecting, they could compare their experience with each step in the approach to identify particularly challenging or novel ideas. Finally, if they had the chance to revisit the problem, by following the approach again, they could improve on their previous approach by applying their new ideas. Overall, this step-by-step approach would act like the scaffolding that supports a building under construction, which is gradually taken down as the building gains more parts. In addition, the step-by-step approach deploys one of the nine ways (segmentation) to reduce cognitive load recommended by [29]. By breaking the complex task of building CNN models for medical image classification into small segments or well-defined steps, the approach could potentially facilitate learning, especially for neurodivergent students [30].

Another specific advantage relates to classroom discussion, as argued by Hollander [31]. The proposed approach can be used as a tool for the discussion where different students read about different steps in the approach and hence structure the discussion based on the approach and minimise digression by asking the students to adhere to the well-defined topics. Moreover, it can also be useful in practical group assignments where an instructor could assign different students to different parts of the approach before asking them to work together to build a CNN-based image classifier. By creating interdependence and judging the group collectively, the instructor would be creating an environment conducive to the jigsaw method of cooperative learning [32]. The instructor could even create multiple groups in this way to promote knowledge diffusion, encouraging this syndicate of groups to devise diverse solutions (multiple scaffolds) to the same problem [33]. In this hypothetical environment, a student would have to complete some tasks independently and creatively (out-of-the-box problem-solving [34], while benefiting from multiple chances to improve on their limited solution through their peers’ perspectives.

Over and above that, one specific benefit of the presented approach is the combinatorial explosion, which is a major challenge in building CNN-based AI applications. There is a wide range of architectures, hyperparameter tuning targets and techniques, and data augmentation techniques. The presented approach is like a checklist to help researchers exhaust the combinations relevant to a medical image classification task. Finally, we have confidence that the organised step-by-step approach will help to classify medical images is increasingly common, and this DL architecture is an essential component of image-classification-based applications. Moreover, this approach brings a common approach that will minimise redundancy, facilitate model comparison, streamline communication, encourage quality control, and promote upscaling. It could also serve as a pedagogical tool. For example, an instructor could base their rubric and grading criteria on our approach. They could use the approach to streamline group work and problem-based learning too.

## Data Availability

All data produced in the present study are available upon reasonable request to the authors.

## Author Contributions

Ahamadullah Lohani: conceptualization, methodology, investigation, software, visualization, resources, and writing and editing; Bhupesh Kumar Mishra: conceptualization, methodology, supervision, validation, resources, and writing and editing; Kenneth Y. Wertheim.: methodology, investigation, validation, and writing and editing; Temitayo Matthew Fagbola: investigation, validation, and writing and editing.

## Data Availability Statement

The data that support the findings of this study are openly available in https://scholar.cu.edu.eg/?q=afahmy/pages/dataset, https://www.kaggle.com/datasets/aryashah2k/breast-ultrasound-images-dataset https://data.mendeley.com/datasets/wmy84gzngw/1https://www.kaggle.com/datasets/vuppalaadithyasairam/ultrasound-breast-images-for-breast-cancer

